# Asthma treatment outcome and factors associated with uncontrolled asthma among adult asthmatic patients in Addis Ababa, Ethiopia

**DOI:** 10.1101/2022.01.18.22269500

**Authors:** Tesfaye Tsegaye, Gebremedhin Beedemariam Gebretekle, Mohammedjud Hassen Ahmed, Tola Bayissa, Bruck Messele Habte

## Abstract

**Background:** Asthma is a major public health challenge in the world resulting in significant health and economic burden. The modifiable and non-modifiable risk factors could have considerable impact on Asthma control and medical care. Our goal was to evaluate the treatment outcome and identify risk factors for poor asthma control among asthmatic patients in Addis Ababa, Ethiopia.

**Method:** A multicentre cross-sectional study using interview and chart review was conducted among asthmatic patients attending ambulatory care of two large public hospitals in Addis Ababa, Ethiopia between March and June 2018. The Global Initiative for Asthma Guideline was used to determine treatment outcomes. The variables of interest were described using descriptive statistics such as frequencies, percentages, mean, and standard deviations. Multivariable logistic regression was used to determine factors associated with uncontrolled asthma. All statistical significance level was determined at p<0.05.

**Result:** A total of 230 asthmatic patients were interviewed. More than half (65.2%) of patients were females and their mean age was 54±15.1 years. Overall, 50.4% of the patients had uncontrolled asthma status. More than two number of trigger factors (AOR=1.88; 95%CI:1.09-2.01), cold weather (AOR=2.11;95%CI: 1.51-2.42), exacerbations of asthma in the last 12 months (AOR=2.01;95%CI:1.39-2.32), moderate persistent asthma (AOR=3.47;95%CI:1.75-5.13), severe persistent asthma (AOR=2.90;95%CI:2.56-3.98) and Salbutamol Puff alone regimen (AOR=2.30;95%CI:2.01-2.76) use were significantly associated with uncontrolled asthma.

**Conclusion:** More than half of asthmatic patients had uncontrolled asthma treatment outcome. This indicates the need to give due attention to asthma patients with uncontrolled status particularly to those with identified risk factors. Health care providers should work in creating patient awareness on appropriate use of their prescribed medications, avoidance of asthma triggering factors for decreasing the progression of the disease and better asthma control.

## Introduction

Asthma is a heterogenous respiratory disease, usually characterized by chronic airway inflammation with variable air flow limitation. (1). It is triggered by dusty environment, cold weather, upper respiratory tract infection, house holde pests, tobacco smoke and strong smell features (2). Asthma is a major cause of morbidity and mortality globally resulting in significant health and economic burden. Globally around 358.2 million people were affected by asthma in 2020 and estimated to affect additional 100 million people by 2025 (3). Every year, about 250,000 people died from asthma which attributes 1.1% of disability-adjusted life years (4,5). It has been documented that if not controlled properly, asthma can affect daily activities and lead to significant physical and socio-economic burden with health related cost (6). Patients with uncontrolled asthma status use the health care service frequently which in turn result in the loss of significant productivity. Compared to those with well-controlled asthma people with uncontrolled asthma bear the largest disease burden and financial burden (7,8). Among the factors reported to aggravate uncontrolled asthma are moderate persistent asthma, severe persistent asthma, exacerbation of asthma, being elderly, persistent exposure to trigger factors, poor knowledge towards asthma care and negative attitude towards inhaled corticosteroids,(6,9,10,11). Recent World Health Organization report showed that there were 6269 or 0.99% total deaths from asthma in Ethiopia. This report further indicated that Ethiopia ranked 61^st^ in number of deaths from among world countries (12). Despite this overarching phenomena few studies have been conducted in Ethiopia to assesss the prevalence of asthma. Studies conducted in Dessie referral hospital, Ethiopia reported a prevalence rate of 10.4% diagnosed asthma (13). Another study reported 29.6% prevalence rate of asthma (14). According to the study conducted in Jimma and Adiss Ababa, Ethiopia, the magnitude of uncontrolled asthma was 64.5% and 53.3% respectively (15,9).

Yet the above mentioned studies have not addressed important variables such as number of trigger factors and type of trigger factors which are thought to be of paramount importance to improve health status of patients with uncontrolled asthma. This study is expected to have important contributions for health professionals involved in the care of patients with asthma in general and those with uncontrolled asthma in particular.

## Methods

### Study area and setting

Addis Ababa, the largest city and political and commercial capital of Ethiopia, has 37 hospitals (12 public, 25 private) and 97 health centers (16). From among these, two public hospitals were selected for this hospital-based cross-sectional study which was conducted from 1 March to 30 June 2018. These two hospitals, i.e., St. Paul’s Hospital Millennium Medical College (St. Paul’s Hospital) and Menelik II Referral Hospital (Menelik II Hospital) were selected for the study due to the high number of patients at follow-up care. St. Paul’s Hospital is one of the largest public teaching hospitals established in 1961 GC. The hospital has 404 beds and serves around 333,752 patients per year, both ambulatory care and hospitalized patients. There were 2692 patients seen in the chest clinic in 2017. Menelik II Hospital was established in 1910 and has 352 beds. The hospital sees more than 206,919 patients every year, with 143 of them requiring asthma treatment follow-up.

### Study population

All asthmatic patients attending ambulatory care of St. Paul’s Hospital and Menelik II Hospital were included for the study based on eligibility criteria. The inclusion criteria were being on anti-asthmatic medications at least for 3 months and age of 18 years or older. Patients with lower and upper respiratory tract infection, chronic obstructive pulmonary disease, emphysema, pulmonary hypertension, congestive heart failure and patients with incomplete chart information were excluded.

### Sample size and sampling techniques

The sample size was calculated using single population proportion formula (17). Considering 53.3% as proportion (p) of uncontrolled asthma (9), with 95% confidence interval and 5% marginal error (d), the sample size was calculated to be 383. However the study population was less than 10,000. Hence, the corrected sample size, using the correction formula was 223. Considering a 5% contingency. The final sample size was calculated to be 234. Participants were recruited from each study settings based on the patient load of the hospitals: 171 from St. Paul’s Hospital and 63 from Menelik II Hospital were allocated proportionally. A consecutive sampling technique was employed to obtain the required number of participants from each hospital.

### Data collection procedures and tools

A structured questionnaire was adopted from Global Initiative for Asthma guidelines (GINA) (6), and by review of other relevant literature (9,15,18,19,20,21) that was used for interviewing selected patients. The pretested structured data collection tool consisted of two parts: (i) information on basic socio-demographic characteristics and (ii) information on asthma symptom control and severity. In addition, patient charts were reviewed to collect information on duration of asthma, comorbidity, concurrent medication(s) and anti-asthmatic medication(s) pattern.

### Statistical analysis

Data were entered into Epi-Data Version 3.1 and then exported to SPSS Version 23.0 for analysis. Descriptive statistics (mean, standard deviation, percentages and frequencies) was used to summarize the data. Multivariable logistic regression analysis was employed to identify factors associated with uncontrolled asthma. A *p-value* of 0.05 or less was considered statistically significant. Asthma treatment outcome was defined based on GINA asthma control tool (6). For this study, the outcome factors were divided into two categories: well-controlled and partially-controlled asthma were designated “controlled”, whereas uncontrolled asthma was considered “uncontrolled”.

### Data quality control

The questionnaire was pre-tested on 5% of the total study participants in the same population outside the study setting in a referral hospital in Addis Ababa for two weeks before the actual data collection time to detect problems and make required revisions. Two days training was given to all data collectors and the first author closely monitored the data collection process. Furthermore, the first author checked collected data for clarity, consistency and completeness on a daily basis.

### Operational definitions

#### Asthma treatment outcome

According to management, asthma severity and control were assessed based on GINA asthma symptom control assessment tool. This tool used to classify treatment outcome status based on daytime symptoms, nighttime symptoms, limitation in activities and rescue medications use (6).

#### Well controlled

It is entitled to an outcome in which patient’s response to asthma symptom control assessment tool like daytime symptoms, nighttime symptoms, limitation in activities and rescue medications used to be none in the past four weeks (6).

#### Partially controlled

This stands for asthma symptom control assessment tool if the study participant’s answer is about one or two “yes’s “(6).

#### Uncontrolled

This stands for asthma symptom control assessment tool in which sample of study participant’s answer is about three or four “yes’s”(6).

#### Treatment outcome

Patients’ response to therapeutic regimen they had been prescribed during clinical follow-up visits based on current clinical subjective findings reported by each study participant. Thus, based on GINA score the overall outcome could be defined into two categories which are controlled and uncontrolled considering GINA score report controlled and uncontrolled asthma. Considering partialy controlled and well controlled as “controlled” asthma status (9).

### Ethical clearance

Ethical clearance was obtained from the Ethical Review Board of School of Pharmacy, Addis Ababa University (protocol number: ERB/ SOP/10/2018). In addition, permission to conduct the study was sought from each hospital. Informed verbal consent was obtained from each study participant after the study was explained to them in detail. Study participants were informed of their right to refuse or discontinue participation at any time and the chance to ask anything about the study. All personal information was kept entirely anonymous and confidential by using of code instead of name, stored in lockable cabinet and password protected.

## Results

### Socio-demographic and Clinical characteristics of patients

A total of 230 participated in the study with a response rate of 98.2%. The mean ±SD age of participants was 54.3 ±15.1 years. The majority of patients were female (65.2%), married (56.5%), and never smokers (85.7%). The mean duration of illness from clinical diagnosis with asthma was 12±9.2 years. About half (47.4%) of the study participants reported that they had at least one time history of asthma exacerbations in the last 12 months and 92 (40%) had moderate persistent asthma as shown in Table 1.

**Table 1:** Socio-demographic and clinical characteristics of patients with asthma at ambulatory care in two public hospitals of Addis Ababa, Ethiopia.

| Characteristics | Frequency (n) | Percentage (%) |
| --- | --- | --- |
| Gender |  |  |
| Male | 80 | 34.8 |
| Female | 150 | 65.2 |
| Age category (in years) |  |  |
| 18-34 | 24 | 10.4 |
| 35-64 | 138 | 60.0 |
| $\geq 65$ | 68 | 29.6 |
| Marital Status |  |  |
| Single | 24 | 10.4 |
| Married | 130 | 56.5 |
| Divorced | 25 | 10.9 |
| Widowed | 51 | 22.2 |
| Residence |  |  |
| Addis Ababa | 193 | 83.9 |
| Out of Addis Ababa | 37 | 16.1 |
| Education |  |  |
| No formal education <sup>a</sup> | 84 | 36.5 |
| Primary education | 68 | 29.6 |
| Secondary education and above | 78 | 33.9 |
| Occupation |  |  |
| Employed | 153 | 66.5 |
| Unemployed | 77 | 33.5 |
| Access of care |  |  |
| Free of payment<sup>b</sup> | 136 | 59.1 |
| Out-of-pocket | 94 | 40.9 |
| Use of biomass fuel |  |  |
| No | 139 | 60.4 |
| Yes | 91 | 39.6 |
| Smoking Status |  |  |
| Never-smoker | 197 | 85.7 |
| Current - smoker | 6 | 2.6 |
| Ex - smoker | 27 | 11.7 |
| Duration of asthma (in years) |  |  |
| < 12 | 148 | 64.3 |
| ≥ 12 | 82 | 35.7 |
| Severity of asthma |  |  |
| Mild intermittent | 50 | 21.7 |
| Mild persistent | 49 | 21.3 |
| Moderate persistent | 92 | 40.0 |
| Severe persistent | 39 | 17.0 |
| Exacerbations in last 12 months |  |  |
| Yes | 109 | 47.4 |
| Comorbidities |  |  |
| Yes | 81 | 35.2 |

<sup>a</sup>: Include illiterates and able to read and write due to informal education like religious teaching
<sup>b</sup>: Include insurance and institutional linkage

### Trigering factors for asthma exacerbations patients with asthma

More than three-fourths (84.8%) of patients reported one or more triggering factors, where cold weather 106 (46.1%) and bad or strong smell 95 (41.3%) were the two leading triggering factors for asthma exacerbations (Table 2).

**Table 2:**
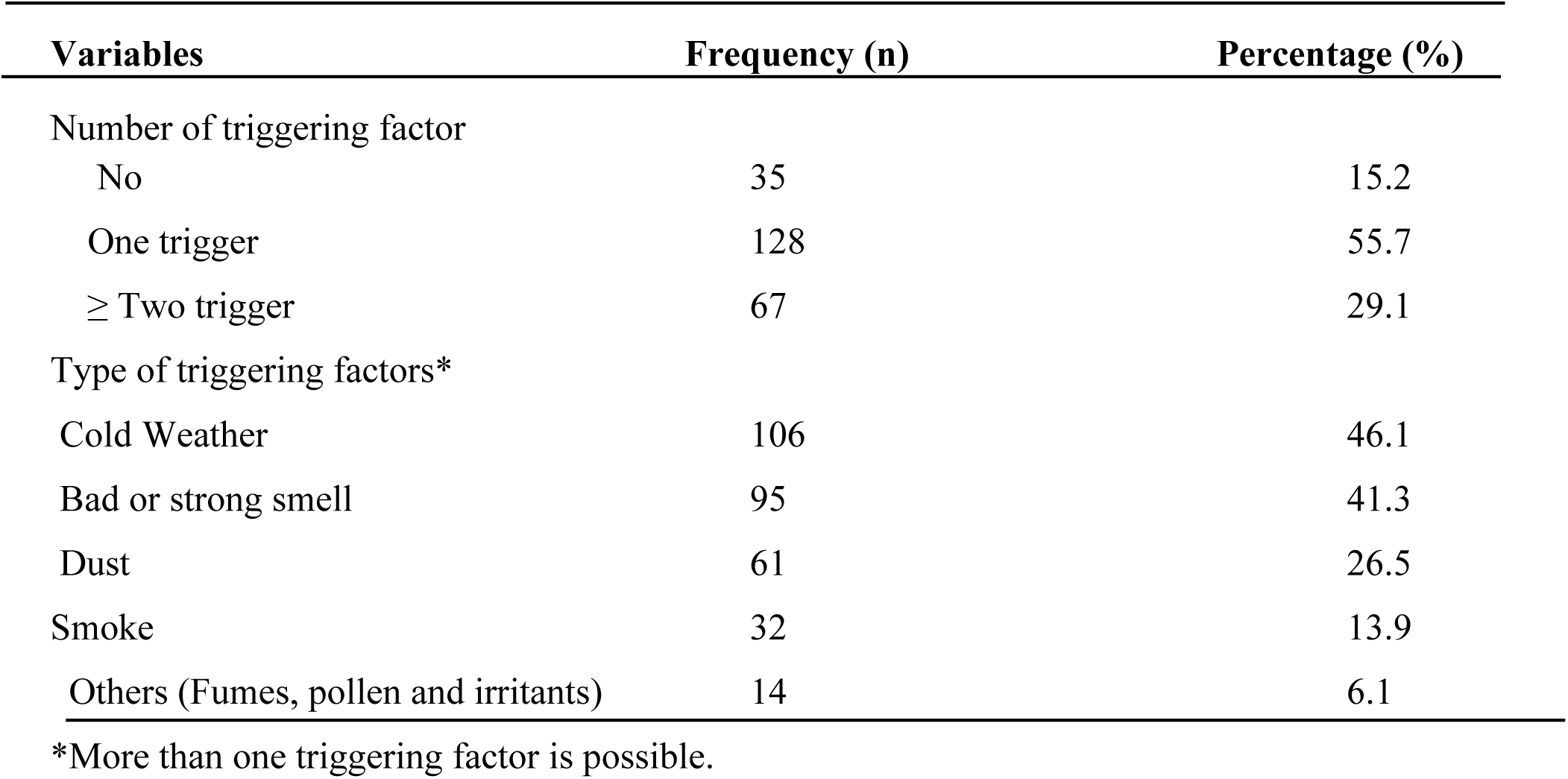
Trigger factors for asthma exacerbations patients with asthma at ambulatory care in public hospitals of Addis Ababa, Ethiopia.

| Variables | Frequency (n) | Percentage (%) |
| --- | --- | --- |
| Number of triggering factor |  |  |
| No | 35 | 15.2 |
| One trigger | 128 | 55.7 |
| ≥ Two trigger | 67 | 29.1 |
| Type of triggering factors* |  |  |
| Cold Weather | 106 | 46.1 |
| Bad or strong smell | 95 | 41.3 |
| Dust | 61 | 26.5 |
| Smoke | 32 | 13.9 |
| Others (Fumes, pollen and irritants) | 14 | 6.1 |

\*More than one triggering factor is possible.

### Anti-asthmatic medication pattern and concurrent medication used by asthmatic patients

Treatment with anti-asthmatic medications was found as alone or combination therapy. Salbutamol puff monotherapy accounted for 54 (23.5%) of the study participants’ treatment while 100 (43.5%) of the patients were using Salbutamol puff + Beclomethasone inhaler. More than one-third (35.2%) of asthma patients had concurrent medications (Table 3).

**Table 3:**
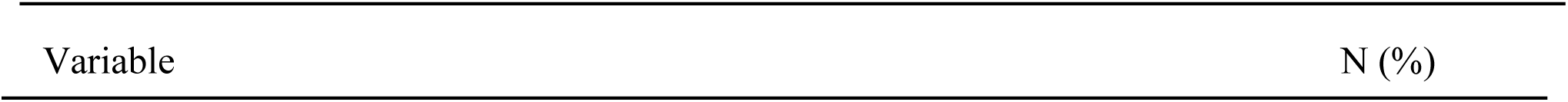
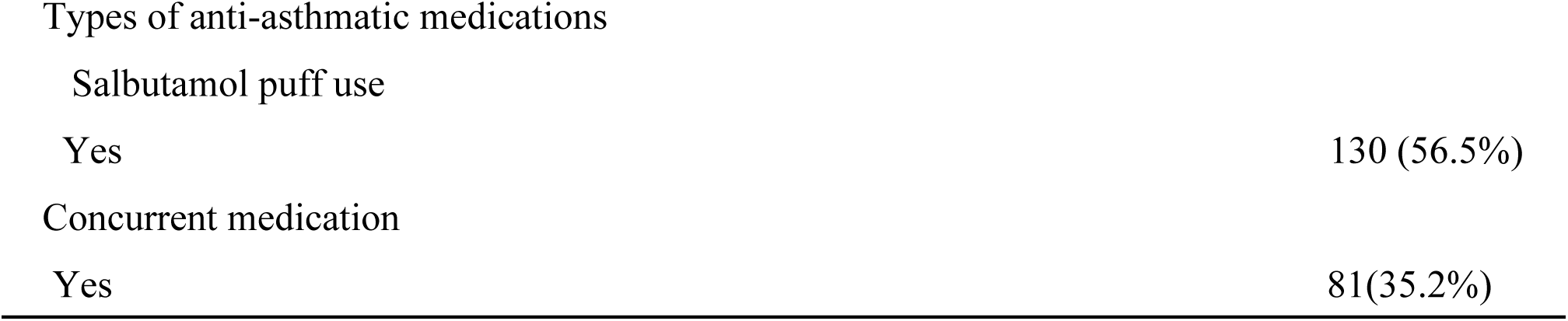
Anti-asthmatic medication pattern and concurrent medication used by asthmatic patients at ambulatory care in two public hospitals of Addis Ababa, Ethiopia.

### Asthma treatment outcome

Over half (50.4%) of patients had uncontrolled asthma while only (22.6%) had well controlled asthma treatment outcome (Table 4).

**Table 4:**
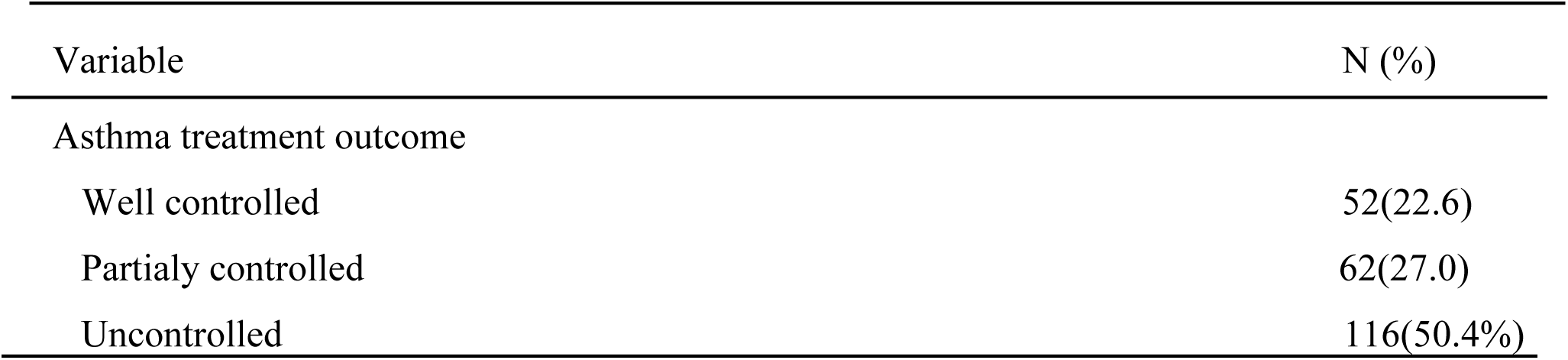
Asthma treatment outcome among asthmatic patients at ambulatory care in two public hospitals of Addis Ababa, Ethiopia.

| Variable | N (%) |
| --- | --- |
| Asthma treatment outcome |  |
| Well controlled | 52(22.6) |
| Partially controlled | 62(27.0) |
| Uncontrolled | 116(50.4%) |

### Risk factors for uncontrolled asthma

Patients with more than two number of trigger factors (AOR=1.88; 95%CI: 1.09-2.01), who had cold weather as triggering factor (AOR=2.11; 95%CI: 1.51-2.42), with asthma exacerbations in the last 12 months (AOR=2.01; 95%CI: 1.39-2.32), with moderate persistent asthma (AOR=3.47; 95%CI : 1.75-5.13) and severe persistent asthma were (AOR=2.90; 95%CI : 2.56-3.98) were more likely to have uncontrolled asthma status than their counterparts. Patients with Salbutamol puff were about 2.30 times more likely to have uncontrolled asthma status than those not taking Salbutamol puff (Table 5).

**Table 5:**
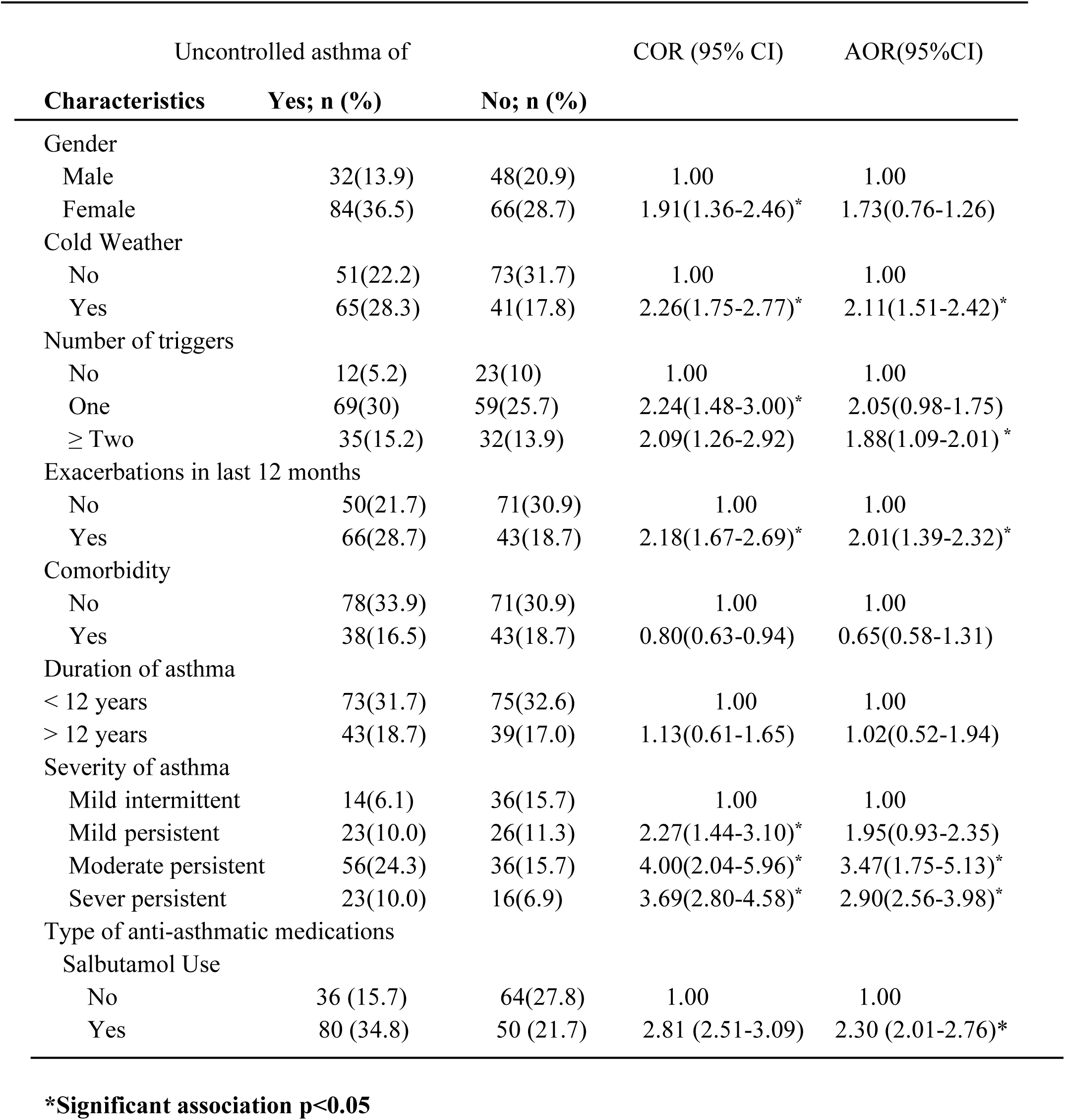
Multivariable logistic regression analysis result of factors associated with uncontrolled asthma among patients with asthma at ambulatory care in public hospitals of Addis Ababa, Ethiopia

## Discussion

This study was conducted to assess the treatment outcome of asthma patients and identify factors associated with uncontrolled asthma among asthmatic patients in Ethiopia. The study showed that more than half of the patients had uncontrolled asthma and several factors such as more than two number of trigger factors, cold weather, exacerbations in the last 12 months, moderate persistent, severe persistent severity of asthma and type of medication, could contribute to the poor treatment outcomes.

Our study revealed that 50.4% of patients had uncontrolled asthma which is comparable to studies conducted in 11 European countries (45%) (22), Vietnam (46%) (23), Italy (51.3%) (18), and Ethiopia (53.3%) (9). This similarity might be due to the use of the same simple screening tool to classify asthma symptom control. This finding was greater than the result reported in China (17.2%) (24) and Saudi Arabia (23.3%) (25). The differences in the level seen might be due to the diagnosis of asthma being made by respiratory physicians, lung function tests done to the patients and the high educational status of study participants in those studies (24,25). In addition to this, the fact that most study participants in those studies have taken health education about asthma disease such as device use technique and behavior related education by their physician and asthma educator might contribute towards the discrepancy (25). However, it was lower than the findings in a study done in South Eastern Nigeria (82.9%) (26) and Jimma, Ethiopia (64.5%) (15). The difference might be explained in part by the fact that patients were found to overestimate their uncontrolled status (26). Compared to the finding from Jimma, Ethiopia, the lower monthly income status of the participants in that study might have led to decreased medications affordability, especially the ICS, which may account towards the difference (15,26).

An asthma standard of management care includes not only the pharmacological treatment but also comprises effective management of triggers as well as risk factors to achieve sustained asthma control (27). Effective and proper utilization of optimized asthma management with anti-asthmatic medication can significantly reduce asthma-related exacerbation (28). Asthma exacerbations in the last 12 months was found to have significant association with uncontrolled asthma, which was consistent with findings from other studies (9,18,20). This might be due to the fact that patients with uncontrolled asthma frequently visit hospital which might increase their chance of acquiring additional disease and exposed to various triggering factors, which in turn result in exacerbations of the symptom (6). In this study moderate and severe persistent asthma was revealed as factors for uncontrolled asthma similar to the study done in Brazil (29) and Jimma, Ethiopia (15). In view of this, it has been documented that patients with moderate and severe persistent asthma were more likely to have uncontrolled asthma (6,29,15)

This study revealed that from pharmaco-therapeutic related factors, patients on Salbutamol puff use alone when compared to those on Salbutamol puff with other controller medicatios were found to have uncontrolled asthma status. The reasons for the observed association might be poor inhaler teqhnique, fear of side effect or dependency and high specific concern towards the controller medications (6), This study finding was in congruent to result reported in previous study (15).

Our study has some limitations. This study being a cross-sectional study doesn’t allow a temporal link to be established between the independent and dependent variables. In addition, the spirometric assessment technique which would have allowed for better measurement of the asthma control level was not used. In spite of these limitations, the study used different assessment tools which helped in acquiring the required data for status of asthma treatment outcome, clinical and behavioral related characteristics of the study.

## Conclusions

The finding of the study indicated that high rates of uncontrolled asthma, more than two number of trigger factors, cold weather, exacerbations in the last 12 months, moderate and severe persistent severity of asthma and Salbutamol use alone were found to be significant predictors to uncontrolled asthma levels. It is therefore recommended that health care providers should work in creating patient awareness of their reliever medications use and avoidance of asthma triggering factors.

## Data Availability

All relevant data are within the manuscript and its Supporting Information files

## Abbreviations

GINA: Global Initiative for Asthma Guideline
ICS: Inhaled Corticosteroids

## Declarations

### Ethical consideration

The study was done in accordance with the principles of the Declaration of Helsinki. Ethical clearance was obtained from Ethical Review Committee of the School of Pharmacy, Addis Ababa University (ERB/SOP/10/10/2018) and SPHMMC (PM23/192) respectively. In addition, the respective heads of the two hospitals permitted the study to be conducted in the clinic. Before questionnaires were administered, informed verbal consent was obtained from each study participant.All other personal information was kept entirely anonymous and confidentiality was secured throughout the study period.

### Consent for publication

Not applicable

### Availability of data and materials

Data used in this analysis are available from the corresponding author on reasonable request.

### Competing interests

The authors declare that they have no competing interests.

### Funding

Addis Ababa university Graduate Programs Office

### Authors’ Contribution

TT participated in almost all parts of the manuscript with support from each of the listed authors. GBG and BMH participated in the protocol writing, study design, review and editing of the manuscript. MHA participated in the data analysis, review and editing the manuscript. TB organized data collection process and analyzed the data, review and editing the manuscript. All authors read and approved the final submission.

## Acknowledgments

We would like to acknowledge the management of St. Paul’s Hospital Millennium Medical College and Menelik II Referral Hospital for facilitating the conduct of this research. Finally, our deepest appreciation goes to all the study participants and data collectors who spent their valuable time for this research work.

## Notes

### Competing Interest Statement

The authors have declared no competing interest.

### Funding Statement

TT received funding from the Graduate Program of Addis Ababa University. The University however had no role in the study design, data collection and analysis, decision to publish or preparation of the manuscript.

### Author Declarations

Ethical Review Committee of the School of Pharmacy, Addis Ababa University (ERB/SOP/10/10/2018) and Saint Paul's Hospital Millennium Medical College (PM23/192)

